# Monitoring COVID-19 transmission risks by RT-PCR tracing of droplets in hospital and living environments

**DOI:** 10.1101/2020.08.22.20179754

**Authors:** Andrea Piana, Maria Eugenia Colucci, Federica Valeriani, Adriano Marcolongo, Giovanni Sotgiu, Cesira Pasquarella, Lory Marika Margarucci, Andrea Petrucca, Gianluca Gianfranceschi, Sergio Babudieri, Pietro Vitali, Giuseppe D’Ermo, Assunta Bizzarro, Flavio De Maio, Matteo Vitali, Antonio Azara, Ferdinando Romano, Maurizio Simmaco, Vincenzo Romano Spica

## Abstract

SARS-CoV-2 environmental contamination occurs through droplets and biological fluids released in the surroundings from patients or asymptomatic carriers. Surfaces and objects contaminated by saliva or nose secretions represent a risk for indirect transmission of COVID-19. We assayed surfaces from hospital and living spaces to identify the presence of viral RNA and the spread of fomites in the environment. Anthropic contamination by droplets and biological fluids was monitored by detecting the microbiota signature using multiplex RT-PCR on selected species and massive sequencing on 16S-amplicons.

A total of 92 samples (flocked swab) were collected from critical areas during the pandemic, including indoor (3 hospitals and 3 public buildings) and outdoor surfaces exposed to anthropic contamination (handles and handrails, playgrounds). Traces of biological fluids were frequently detected in spaces open to the public and on objects that are touched with the hands (>80%). However, viral RNA was not detected in hospital wards or other indoor and outdoor surfaces either in the air system of a COVID-hospital, but only in the surroundings of an infected patient, in consistent association with droplets traces and fomites. Handled objects accumulated the highest level of multiple contaminations by saliva, nose secretions and faecal traces, further supporting the priority role of handwashing in prevention.

In conclusion, anthropic contamination by droplets and biological fluids is widespread in spaces open to the public and can be traced by RT-PCR. Monitoring fomites can support evaluation of indirect transmission risks for Coronavirus or other flu-like viruses in the environment.

**Importance:** Several studies searched for SARS-CoV-2 in the environment because saliva and nasopharyngeal droplets can land on objects and surfaces creating fomites. However, the ideal indicator would be the detection of the biofluid. This approach was not yet considered, but follows a traditional principle in hygiene, using indicators rather than pathogens. We searched for viral RNA but also for droplets on surfaces at risk. For the first time, we propose to monitor droplets thorugh their microbiota, by RT-PCR or NGS.

Even if performed during the pandemic, SARS-CoV-2 wasn’t largely spread on surfaces, unless in proximity of an infectious patient. However, anthropic contamination was frequently at high level, suggesting a putative marker for indirect transmission and risk assessment. Moreover, all SARS-CoV-2-contaminated surfaces showed the droplets’ microbiota.

Fomites detection may have an impact on public health, supporting prevention of indirect transmission also for other communicable diseases such as Flu and Flu-like infections.

**GRAPHICAL ABSTRACT:** 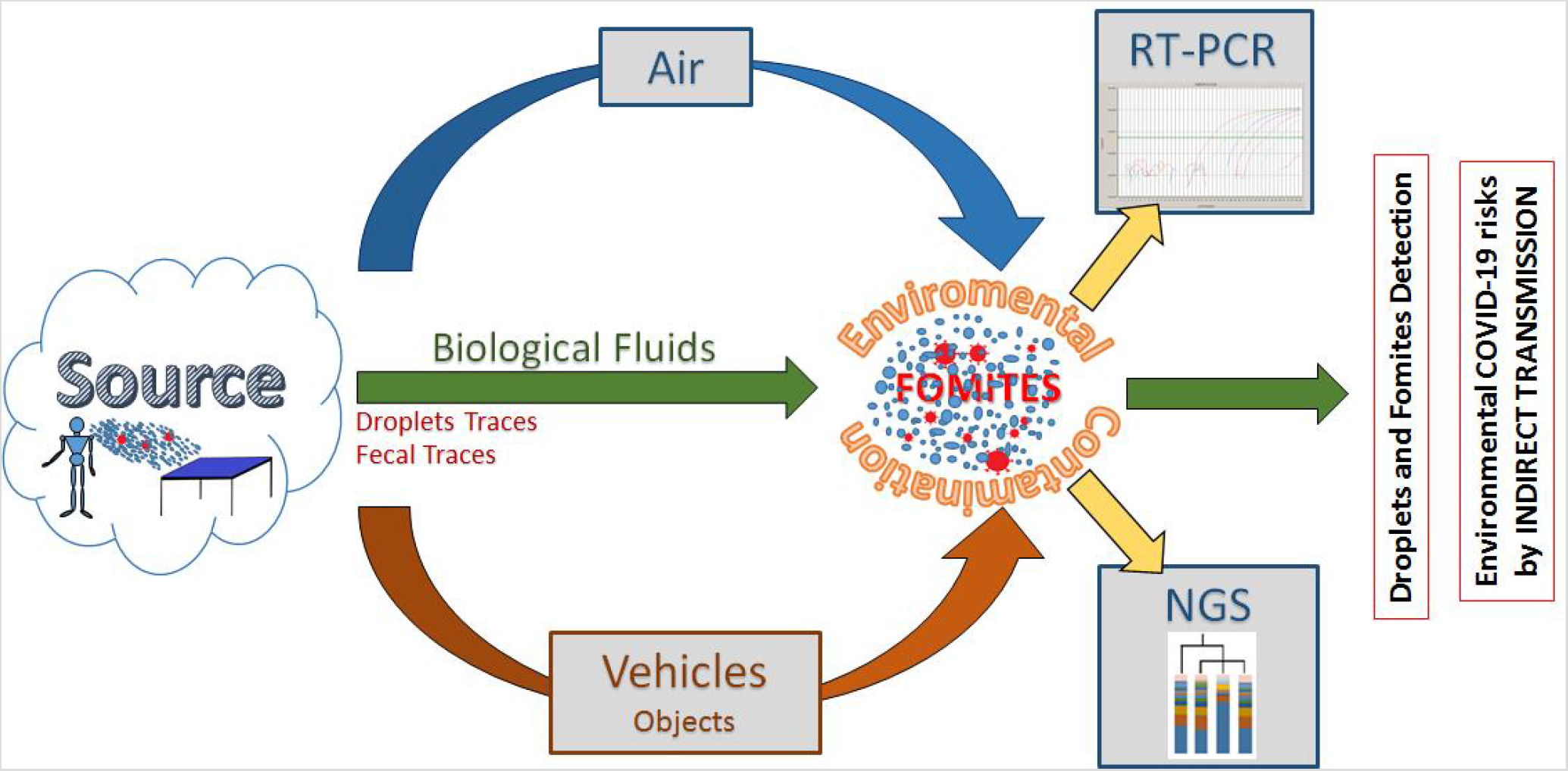

## 1 Introduction

The ongoing pandemic of coronavirus disease 2019 (COVID-19) is thought to have spread mainly through a direct route of transmission: by person to person close contact and inhalation of virus-laden liquid droplets (WHO, 2020; Stadnytskyi et al., 2020; Somsen et al., 2020; Liu et al., 2020; NCIRD, 2020; Asadi et al, 2020). However, it is also recognised that Severe Acute Respiratory Syndrome Coronavirus 2 (SARS-CoV-2) can follow an indirect route of transmission through environmental contamination of objects and surfaces, being carried by biological fluids such as saliva, nose secretions, and feces, in which viral RNA was consistently detected (Van Doremalen, et al., 2020; Chia et al., 2020; Guo, et al., 2020; Cheng et al., 2020; Ong et al., 2020; Lv et al., 2020; Razzini et al., 2020; Li et al., 2020; Chen et al., 2020; Foladori et al., 2020). Respiratory droplets (aerodynamic diameter ranging between > 5 and 10 µm) and droplet nuclei or aerosols (≤ 5 µm) from a patient or an asymptomatic subject can reach directly the mouth, nose or eyes of a susceptible person, but can also land in the surroundings, creating contaminated surfaces, namely fomites (WHO, 2014; Wei et al., 2020; Otter et al., 2016; Al Huraimel et al., 2020). The risk of an indirect transmission through contaminated surfaces is related to the capability of the coronavirus to survive on different matrices under different conditions, persisting from hours to days, especially in indoor environments (Eslami and Jalili. 2020; Kampf et al., 2020; Hung et al., 2020). The question of evaluating the level of environmental contamination is important not only during outbreaks and the epidemic peak, but also during the transition phases, when asymptomatic infected carriers may release the virus in the environment by talking, coughing, sneezing, touching objects and consequently spreading contaminated biofluids (including saliva and nose secretions, faecal traces or droplets) (Kampf et al., 2020; Hung et al., 2020; Wu et al., 2020). This issue has already been considered in higher-risk environments such as hospitals and public areas, where SARS-CoV-2 was clearly detected in several studies, even when surveillance and sanitation were accurately performed (Wu et al., 2020; Wang et al. 2020).

Finally, when approaching the question of the presence of SARS-CoV-2 in the environment, we need to consider that the pathogen is conveyed by biological fluids. Therefore, the possibility of detecting fomites and biological fluids in the environment becomes both a potential marker of hygiene levels, and also a candidate indicator for indirect COVID-19 transmission risks. Starting from this working hypothesis, we surveyed hospitals and public buildings not simply for the presence of SARS-CoV-2, but also for the presence of droplets, fomites and anthropic contaminations, by searching traces of the microbiota signature of their own biological fluids of origin. To address this issue, a dedicated set of primers and probes was combined to detect different biological fluids based on multiplex reactions in real-time PCR (RT-PCR), following a strategy initially developed for forensic studies and hospital hygiene (Giampaoli et al., 2012; Giampaoli et al., 2014; Valeriani et al., 2016; Valeriani et al., 2018a; Yao et al., 2020). Thus, a same general approach based on RT-PCR was considered to test both the viral RNA by standard procedures and the DNA from fomites by microbiota signature. The general principle is based on the amplification of genes from at least one representative bacterial component of the biological fluid, e.g. amplifying bacterial genes from *E. salivarius* and *S. mutans* for targeting saliva, or *Corynebacterium* for nose secretions, or *E. faecalis* and *Bacteroides* for evaluating the presence of faecal traces, as previously shown (Esberg et al., 2020; Liu et al., 2020; Charles et al., 2019; Proctor et al., 2019; Lloyd-Price, J et al., 2016). This approach can be confirmed by next generation sequencing (NGS), analysing the whole microflora DNA (mfDNA) as sampled with environmental swabs on indoor and outdoor surfaces (Slatko et al., 2018; Valeriani et al., 2018b; Valeriani et al., 2018c; Valeriani et al., 2019; Mucci et al., 2020).

The overall aim of this study was to search for both SARS-CoV-2 and fomites in hospitals and public buildings, to evaluate the possibility of monitoring by RT-PCR fomites and biofluid contamination, as a novel indicator of hygiene as well as a candidate marker for indirect transmission risks for COVID-19.

## 2. Materials and Methods

### 2.1. Sampling and experimental design

Surfaces at risk for the presence of biological fluids and the transmission of COVID-19 were sampled from different settings, both in indoor and outdoor areas. Environmental samples (n = 94) were collected after the epidemic peak in Italy (May-June 2020). We sampled indoor surfaces from three COVID-reference hospitals in three Italian regions (Parma, Emilia-Romagna; Sassari, Sardinia; Rome, Lazio); buildings open to public use (1 office, 1 fast food, 1 church); outdoor areas; and used handkerchiefs with nasopharyngeal secretions. Samples were tested for SARS-CoV-2 RNA, whereas anthropic contamination was assessed searching for biological fluids of nose, mouth, gut through their microbiota traces by RT-PCR and/or NGS (Table 1).

**Table 1.** Environmental sampling: dataset of collected samples and testing. Indoor surfaces were sampled from different hospitals (n = 3), buildings of public use (1 office, 1 Fast food, 1 church), outdoor areas (n = 16), nose-oropharyngeal secretions (n = 4) and tested for presence of SARS-CoV-2 and for anthropic contamination by testing the presence of microbiota traces of biological fluids (Nose, Saliva, Feces), by Real Time PCR (RT-PCR) and/or Next Generation Sequencing (NGS). For each type of test, analysis was performed in all (+), no one (−) or some (±) of the collected samples.

| *Classification* | *Samples* | **Dataset**<br>**(N=94)** | **Description** | **Nucleic Acid Testing** |  |  |
| --- | --- | --- | --- | --- | --- | --- |
| *Surface source* |  |  |  | SARS-CoV-2 | Biofluid (RT-PCR) | Microbiota (NGS) |
| **Indoor** | **Hospital** | **49** | Floor (n=7) | + | + | + |
|  |  |  | Bedside table (n=3) | + | + | + |
|  |  |  | Door handle (n=3) | + | + | + |
|  |  |  | Call button (n=1) | + | + | + |
|  |  |  | Table (n=1) | + | + | + |
|  |  |  | Chair (n=2) | + | + | + |
|  |  |  | Back of the bed (n=3) | + | + | + |
|  |  |  | Side of bed (n=4) | + | + | + |
|  |  |  | Bottom of the bed (n=1) | + | + | + |
|  |  |  | Wall behind the bed head (n=1) | + | + | + |
|  |  |  | Bed sheets (n=1) | + | + | + |
|  |  |  | Pillow (n=2) | + | + | + |
|  |  |  | Stethoscope (n=1) | + | + | + |
|  |  |  | Wheelchair head (n=1) | + | + | + |
|  |  |  | Toilet board (n=1) | + | + | + |
|  |  |  | Toilet flush button (n=1) | + | + | + |
|  |  |  | Sink faucet (n=1) | + | + | + |
|  |  |  | Air circulation system (15) | + | - | - |
|  | **Public Building** | **25** | Door handle (n=2) | ± | + | + |
|  |  |  | Toilet (n=2) | + | + | + |
|  |  |  | Pews (n=4) | - | + | + |
|  |  |  | Floor (n=6) | ± | + | + |
|  |  |  | Toilet wall tiles (n=3) | ± | + | + |
|  |  |  | Office phone (n=1) | + | + | + |
|  |  |  | Computer keyboard (n=2) | + | + | + |
|  |  |  | Air circulation system (n=5) | - | + | + |
| **Outdoor** |  | **16** | Handrail (n=1) | + | + | + |
|  |  |  | Grip shared e-scooter (n=1) | + | + | + |
|  |  |  | Bus stop bench (n=1) | + | + | + |
|  |  |  | Coffee dispenser button (n=1) | + | + | + |
|  |  |  | External door handle (n=2) | ± | + | + |
|  |  |  | Playground (n=10) | - | ± | + |
| **Human** |  | **4** | Droplets/biofluid (n=4) | ± | + | + |

### 2.2. Sampling collection

Surface sampling was carried out after their use and prolonged exposure to human presence (>4h). Following standard protocols, FLOGSwabs and CITOSSWAB were used and soaked into a buffer solution in a volume of 400 µl of UTM-RM transport medium (Copan Diagnostics Inc., Murrieta, CA, USA). The nasopharyngeal secretions were collected on handkerchiefs with swabs (4N6FLOQSwabs, Copan Diagnostics Inc., Murrieta, CA, USA). All specimens were refrigerated at 4°C if testing could postpone in the following days.

### 2.3. SARS-CoV-2 detection

All samples in UTM were heat inactivated at 56°C for 5 minutes to reduce the risk of accidental transmission of SARS-CoV-2 to laboratory personnel. Nucleic acids were purified and extracted using the eMag automated nucleic acid sample extraction system (bioMérieux, Marcy l’Etoile, France). Briefly, total nucleic acids were extracted from UTM using an input sample volume of 200 ml into 2,000 ml of easyMag lysis buffer using B protocol to a final eluted volume of purified nucleic acids of 50 ml. TaqPath 1-step reverse transcriptase quantitative PCR (RT-qPCR) master mix (Life Technologies, Frederick, MD) and the 2019-nCoV CDC EUA kit (Integrated DNA Technologies, Coralville, IA) were used for target detection (*CDC, 2020*). Molecular detection of SARS-CoV-2 RNA was carried out by rRT-PCR, using primers and probes related to the E gene, with a detection limit of 5.2 copies of RNA/reaction (Corman V. 2019). Samples were analyzed in Sassari and Parma with the Allplex 2019-nCoV assay (Seegene, Seoul, South Korea) and in Rome with the Detection Kit for 2019 Novel Coronavirus (2019-nCoV) RNA (PCR-Fluorescence Probing) (Daan Gene Co., Ltd of Sun Yat-University, Guangzhou, Guandong, China) for the confirmation of the results. The Allplex 2019-nCoV assay was designed for amplifying three viral targets: the E gene (subgenus Sarbecovirus), the N, and the RdRP genes (Farfour, 2020). The Detection Kit for 2019 Novel Coronavirus (2019-nCoV) RNA (PCR-Fluorescence Probing) is a CE-marked RT-PCR assay which simultaneously detects the viral nucleocapsid N and Orf1ab genes. Afterward, 5 µl of eluted RNA samples in a total volume of 25 µl were RT-PCR amplified on Biorad CFX96 real-time system. In each round of extraction and amplification, positive and negative control samples (supplied by the manufacturer) were included. The interpretation criteria were the following: 1) positive signals detected in ORF1ab and N genes with a cycle threshold (Ct) values ≤40 were considered positive for SARS-CoV-2 RNA; 2) positive signals in only one gene (N or Orf1ab) with a Ct values ≤40 were considered inconclusive; and 3) no fluorescent signals or over the 40th Ct in ORF1ab and N genes were considered not specific and reported as negative for SARS-CoV-2 RNA. The declared LoD is 500 RNA copies/ml.

### 2.4. DNA extraction

An aliquot of COPAN UTM-RM transport medium (about 300 μL) was centrifugated at 16000 g for 10 minutes and the pellet was manually disaggregated with a pestle after adding glass beads (Sigma-Aldrich, St Louis, MO) and lysed in 200 μL lysozyme solution, RNase A treated, and proteinase K digested according to the GenElute Bacterial Genomic DNA Kit (Sigma Aldrich, St Louis, USA). Finally, DNA elution was performed in 60 μL elution solution (10 mM tris(hydroxymethyl)aminomethane-hydrochloride and 0.5mM ethylene diamine tetra acetic acid, pH 9.0). For pharyngeal biofluids and fomites samples, each swab was inserted into the semipermeable NAO Baskets and broken inside at the breakpoint. Approximately 200 μL lysozyme solution (20mg/mL Lisozima, 20 mM tris[hydroxymethyl]aminomethanehydrochloride at pH 8, 2mM ethylenediaminetetraacetic acid, and 1.2%TritonX-100; Sigma Aldrich, St Louis, USA) were added into the NAO Baskets and incubated a 37°C for 30 minutes. Then, 20 μL proteinase K and 400 μL buffer AL were added and the sample was centrifuged at 10,000 × g for 1 minute, allowing the elution of the digestion solution. After incubation at 56°C for 10 minutes and addition of 400 μL ethanol, the washing step and DNA purification were performed in accordance with the manufacturer’s instructions. DNA elution was completed in 60 μL elution solution (10 mM tris[hydroxymethyl]aminomethane-hydrochloride and 0.5 mM ethylenediaminetetraacetic acid at pH 9.0), as previously described (Cianfanelli et al., 2017; Valeriani et al., 2017; Valeriani et al., 2018c;).

### 2.5. Analysis of mfDNA by multiplex real-time PCR and data interpretation

Amplifications were combined in 4 multiplex reactions: mix Skin, for the identification of *Staphylococcus aureus* and *Staphylococcus epidermidis*; mix Nasopharynx for *Propionibacterium spp*. and *Corynebacterium spp*.; Mix oralpharinx for *Streptococcus salivarius* and *Streptococcus mutans*; mix feces for *Enterococcus spp* and *Bacteroides vulgatus* (probes were labeled FAM/VIC/HEX, with the BHQ-1quencher). Primers for different bacterial indicators and optimized reaction conditions were already established, as previously described (Elshi et al., 2000; Giampaoli et al., 2012; Chung et al., 2016; Valeriani et al. 2018a; Byrd et al., 2018; Liu, Q. 2020). Briefly, amplifications were performed in a volume of 25 μL, of which 12.5 μL JumpStart Taq ReadyMix for Quantitative PCR (Sigma Aldrich, St. Louis, MO), containing 900nM forward and reverse primers, and 250 nM of each probe. For each mix, samples were tested in triplicate. The amplifications were performed using Bio-RadCFX96(Bio-Rad,Hercules, CA) programmed for 10 minutes at 95°C and 40 cycles of 15 seconds at 95°C and 1 minute at 60°C. For each sample 11 μL template reaction was amplified. The PCR output was expressed as cycle threshold (CT). Positive samples were those where ≥1 positive indicator (CT ≤35) was found in at least 2 mixes. Conversely, a microbial indicator was considered negative when over the CT ≥39 threshold.

### 2.5. 16S rDNA Amplicon sequencing analysis

Libraries for NGS were prepared according to the 16S Metagenomic Sequencing Library Preparation Guide (part# 15044223 rev A; Illumina, San Diego, CA, USA). The PCR amplicons were obtained using Ba27F and Ba338R primers containing overhang adapters, as previously described (40, 41). Tagged PCR products were generated using primer pairs with unique barcodes through two-step PCR. In this strategy, target primers containing overhang adapters were used in the first PCR reaction to amplify the target gene, that product was then used in the second PCR using primers-containing barcodes. Each amplification reaction had a total volume of 25 μL, containing 12.5 μL of KAPA HiFi Hot Start Ready Mix (Roche, Pleasanton, CA, USA), 5 μL of each primer (1 μM), and 2 μL of template DNA. Reactions were carried out on a Techne®TC-PLUS thermocycler (VWR International, LLC, Radnor, PA, USA). Following amplification, 5 μL of PCR product from each reaction was used for agarose gel (1%) electrophoresis to confirm amplification. The final concentration of cleaned DNA amplicon was determined using the Qubit PicoGreen dsDNA BR assay kit (Invitrogen, Grand Island, NY, USA) and validated on a Bioanalyzer DNA 1000 chip (Agilent, Santa Clara, CA, USA). Libraries were prepared using the MiSeq Reagent Kit Preparation Guide (Illumina, San Diego, CA, USA). Raw sequence data was processed using an in-house pipeline that was built on the Galaxy platform and incorporated various software tools to evaluate the quality of the raw sequence data (FASTA/Q Information tools, Mothur). All datasets were rigorously screened to remove low-quality reads (short reads > 200 nt, zero-ambiguous sequences). Demultiplexing was performed to remove PhiX sequences and sort sequences; moreover, to minimize sequencing errors and ensure sequence quality, the reads were trimmed based on the sequence quality score using Btrim (an average quality score of 30 from the ends, and remove reads that are less any 200 bp after end-trimming) (Kong, 2011). OTUs (operational taxonomic units) were clustered at a 97% similarity level, and final OTUs were generated based on the clustering results, and taxonomic annotations of individual OTUs were based on representative sequences using RDP’s 16S Classifier 2.5. Observed OTUs were defined as observed species. A level of 97% sequence identity is often chosen as representative of a species and 95% for a genus. The sequence reads were analyzed, also, in the cloud environment BaseSpace through the 16S Metagenomics app (version 1.0.1; Illumina®): the taxonomic database used was the Illumina-curated version (May 2013 release of the Greengenes Consortium Database) (Wang et al., 2013).

### 2.6. Statistical Analysis

Relative abundances of community members were determined with rarefied data and summarized at each taxonomic level. The proportion of the microbiome at each taxonomic rank, such as phylum, order, class, family, and genus, was determined using the RDP classifier and the Greengenes Database. Alpha and beta diversity were calculated using EstimateS software at a level of 97% sequence similarity. Regarding alpha diversity, the Shannon index and equitability index at the species level were computed (Colwell et al., 2012; Magurran, 2016). Principal component analysis (PCA) was performed using the METAGENassist platform and and R (version 3.1.3, www.R-project.org) with packages “ggplot2”, “ape”, “psych” and “vegan” (Arndt et al., 2012). Multivariate analysis, the PCA, and partial least square-discriminant analysis (PLS-DA) were performed in order to investigate the dissimilarity between groups. Feature selection was performed using PLS-DA and 10-fold cross validation to tune algorithm parameters and to check model validity. Dendrogram and clustering analysis were based on the Euclidean distance and Ward’s method.

## 3. Results and discussion

### 3.1 Detection of fomites by RT-PCR

Anthropic contamination by droplets and biofluids was detected on different environmental surfaces by RT-PCR (Table 2). The presence of fomites appeared largely diffused in indoor and outdoor areas exposed to human crowding or frequently touched with hands. Floors and walls were less contaminated than handles or buttons. Droplets DNA traces were detected in most of surfaces and almost 10% of sampled points displayed a multiple contamination from different biological fluids. Correlation between selected bacterial species and biological fluids in droplets and fomites was confirmed (p-value < 0.01) (Table 3), supporting the effectiveness of the approach. The combined action of different markers is synergic (Figure 1), allowing a reliable identification of droplets and fomites. Indoor and outdoor samples showed the presence of traces from one or more human biofluids, although with different intensity. Being a quantitative approach, indeed, RT-PCR can provide not only a qualitative output for the presence of droplets traces, but also an indication on the contamination level, allowing to set different thresholds or report CT data in the form of genomic units. However, the definition of a more precise quantitative interpretation of the multiplex amplification output would require a larger exploitation of the method, based on different environments, monitoring purposes or expected hygiene levels. The procedure to extract nucleic acid and detect anthropic contamination from environmental swabs could be automatize, likewise already done for testing SARS-CoV-2 from human swabs; therefore, detection of fomites by RT-PCR seems a feasible and promising approach even on a larger scale.

**Table 2:**
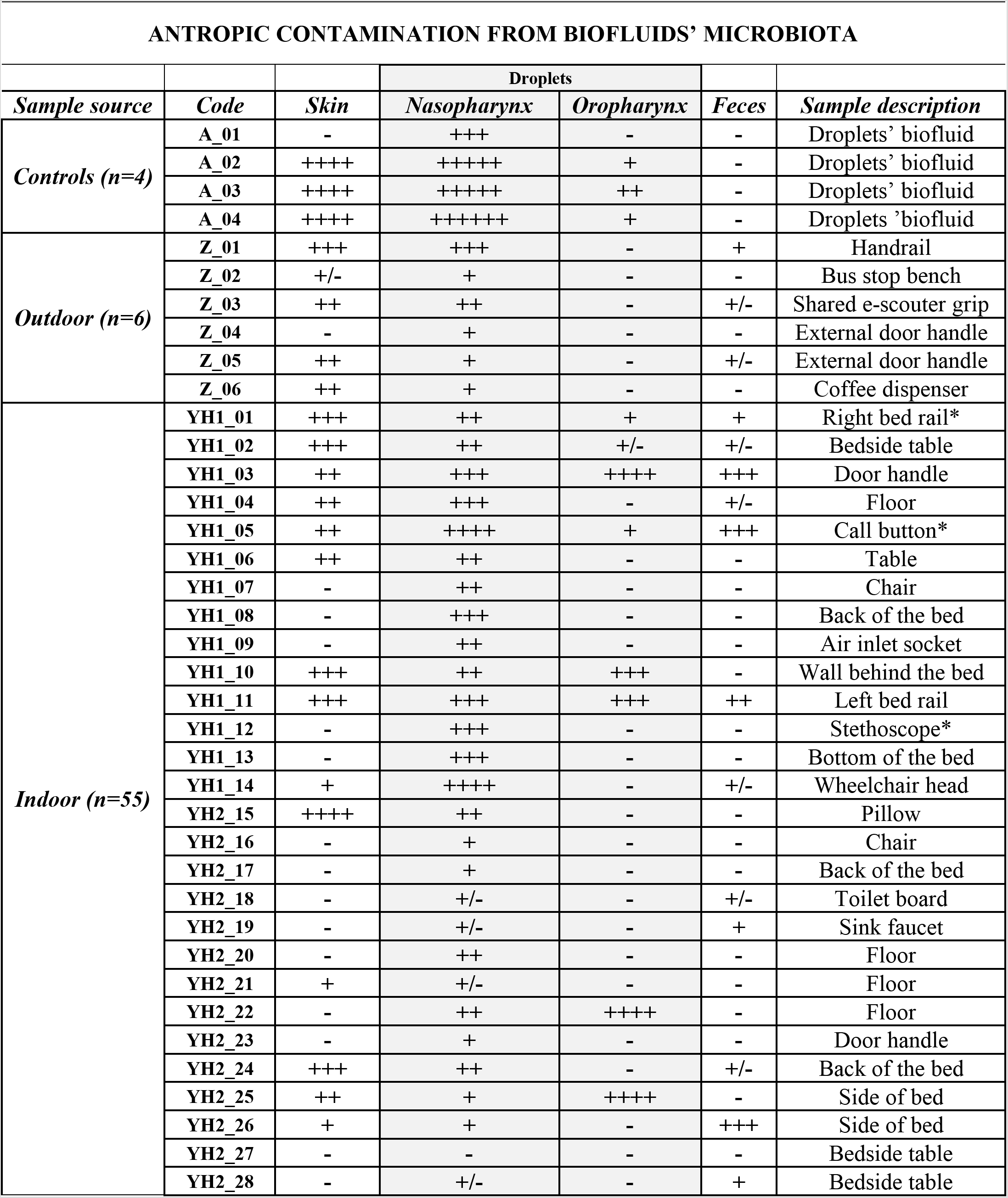
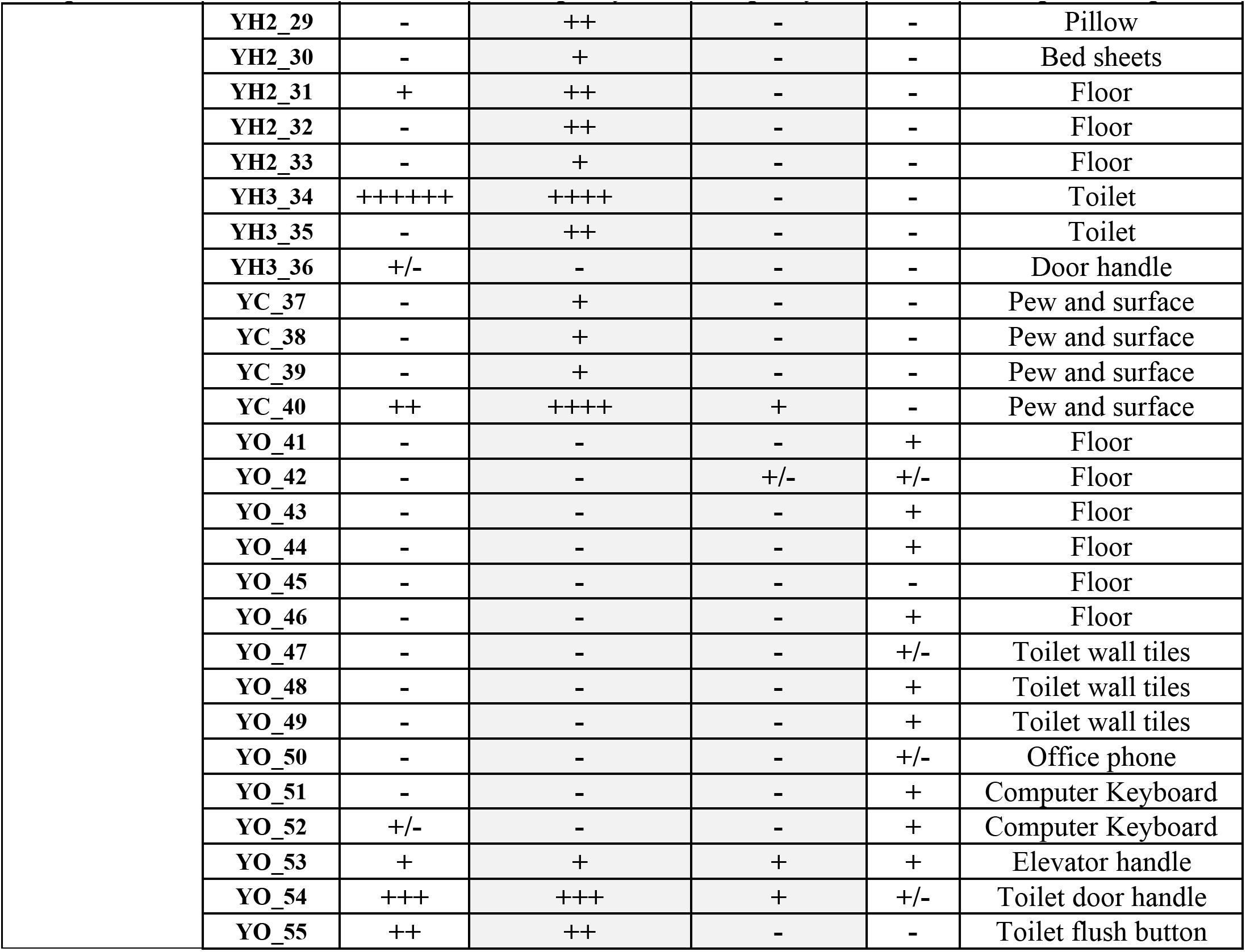
Anthropic contamination by Real Time PCR. The cumulative output of indicators for each anthropic contamination is shown, including the description of each sample. For each indicator: “+++” Positive: Ct < 20; “++” Positive: Ct 20-30; “+” Positive Ct 30-35; “+/−” Doubt: Ct 36-38; “-” Negative Ct > 39. For *E. faecalis*: “++” Positive: Ct < 20; “+” Positive Ct 21-29; “+/−” Doubt: Ct 30–35; “-” Negative Ct > 36. The asterisk (*) shows the sampling points where SARS-CoV-2 RNA was detected. Sample reading code: A: anthropic, Z: outdoor; Y: indoor (H: hospital; C: church; O: office and restaurant).

**Table 3.**
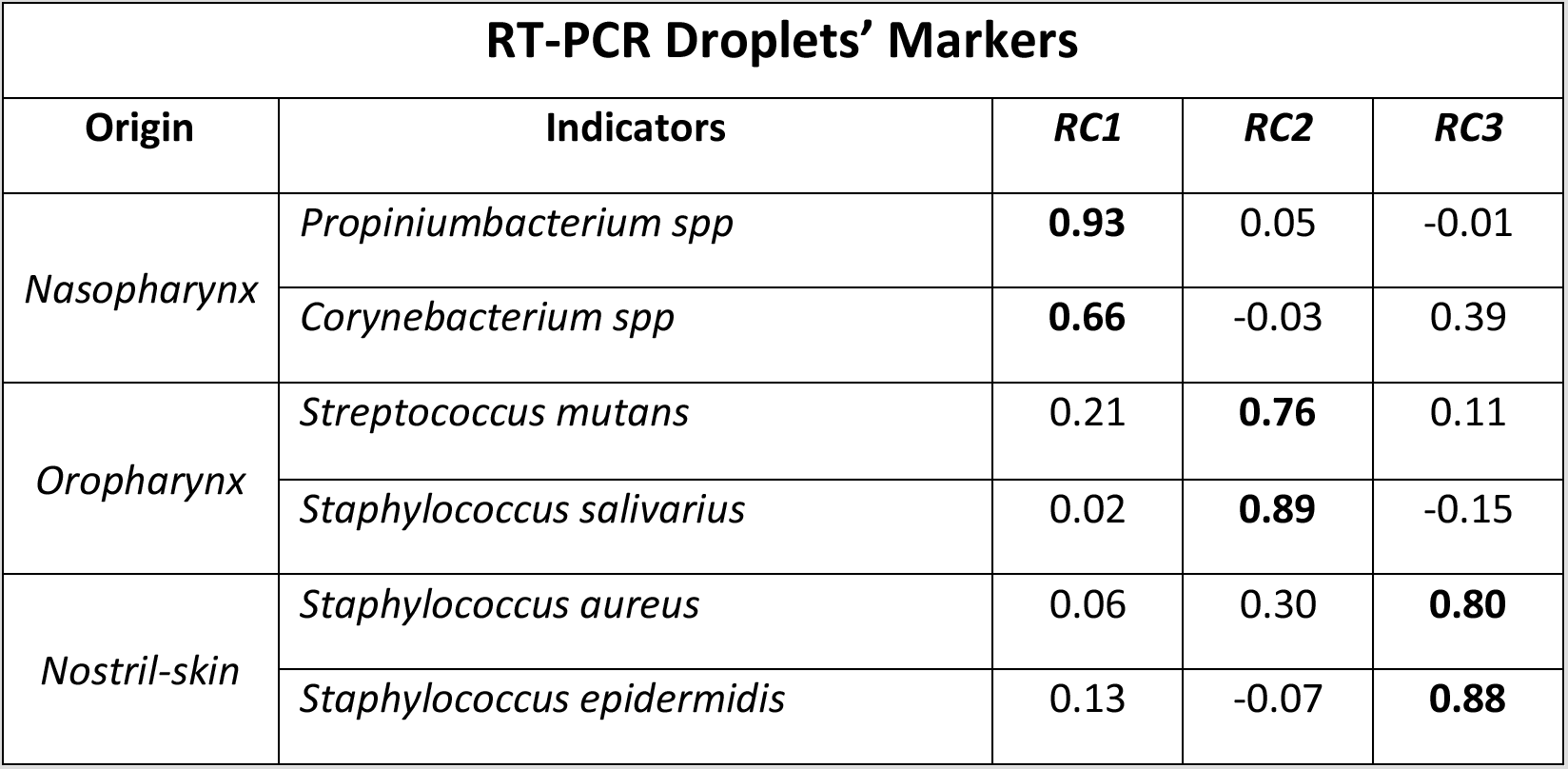
Primary indicators for droplets. Pattern matrix for principal component analysis nasopharynx selected indicators *(Propiniumbacterium spp., Corynebacterium spp.)*, oropharynx *(Streptococcus mutans, Staphylococcus salivarius*), of nostril-skin (*Staphylococcus aureus, Staphylococcus epidermidis)*. The higher correlations for each component (RC) are shown in bold (p< 0.01). The table reported variable loading on the rotation matrix.

**Figure 1.**
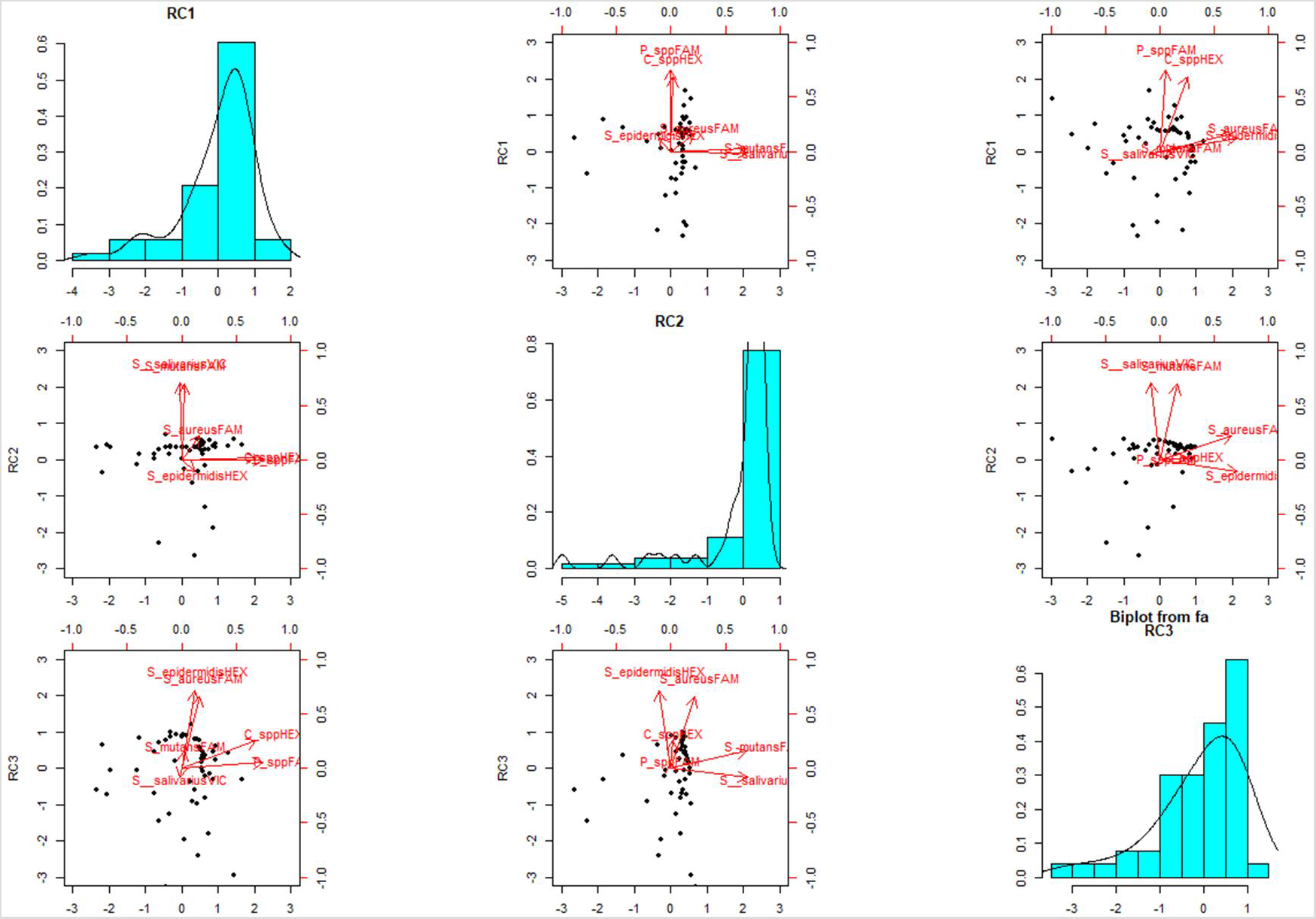
Droplets’ components’ distribution. Principal Component Analysis biplots Component 1 versus Component 2, Component 1 versus Component 3 and Component 2 versus Component 3. The first, second, and third component explain 28%, 27%, and 27% of the variability observed, respectively. The role of the bacterial indicators within the different components is summarized by vectors within the scatter graphs.

### 3.2 SARS-CoV-2 in hospitals and public places

SARS-CoV-2 was not detected in most of the sampling points, both indoor and outdoor. It was not found in all 15 sampling points within the air system of a COVID-19 hospital. SARS-CoV-2 RNA was detected only in one room where an infected patient was hospitalized and only in those samples collected near to the patient (one on the bed rail and one on the surface of the call button). The stethoscope used on the patient was positive, too. The low frequency (< 4%) of positive samples in comparison with other studies (20-30%) can be associated to differences in the sampling strategy or to a lower sensitivity of the method (Liu, Y., 2020); moreover, it depends on the epidemiological scenario when the study was carried out, at time when reopening of activities was carefully surveilled and preventive measures were strictly enforced (Pasquarella, 2020; Di Maria, 2020; Veronesi, 2020; Van Doremalen, et al., 2020; Chia et al., 2020; Guo, et al., 2020; Cheng et al., 2020; Ong et al., 2020; Lv et al., 2020; Razzini et al., 2020; Li et al., 2020; Chen et al., 2020; Foladori et al., 2020; Wei et al., 2020). From our findings, it seems that the environmental spread of SARS-CoV-2 was not widely diffused, with the only exception of the surfaces near a hospitalized infected patient.

### 3.3 NGS analysis

The microbial signature obtained by RT-PCR was confirmed by NGS and all selected bacterial indicators were also included within the microbiota identified by high-throughput sequencing and bioinformatic analysis. RT-PCR and NGS characterized environmental samples based on their contamination patterns: by CT analysis on selected marker genes or by reads count on all 16S amplicon sequences, respectively (Figures 2 and 3). DNA test can be easily performed within one day on any real time apparatus, whereas NGS within one week adapting the laboratory protocol to the high throughput sequencer and accomplishing the required bioinformatic evaluation on the obtained data. However, NGS provides the 16S rDNA sequences of all the bacterial species in the sample, so that any anthropic contamination can represent just only a minority of the resident or acquired microflora on a surface. Therefore, each selected indicator specifically amplified by RT-PCR was observed by NGS, but only as a subcomponent between others (about 200-1000 species for each sample), including the unknown species (about 5-10%). Mean values ranged from 0.24% (*Bacteroides*) to 5.78% (*Corynebacterium*). For example, *Corynebacterium* showed lower values (< 1%) in environmental samples exposed to multiple sources of microbial pollutions, whereas the highest values (35-80%) were observe in environmental swabs collected from human nasopharyngeal secretions, further confirming the specific role of that marker within the microbial signature of the biological fluid. Nevertheless, both methods provided similar dendrograms. The correspondence between RT-PCR and NGS was reliable: two independent SARS-CoV-2 positive samples (YH1_01 and YH1_05) collected from same patient surroundings closely gathered, whereas the sample collected from the stethoscope used on the same patient (YH1_12) segregated at a different distance, in both dendrograms. Indeed, the anthropic contaminations present on the right side of the bed (YH1_01) and on the call bottom (YH1_05) showed a similar biodiversity pattern (as also shown by the Shannon index 2.602 and 2.893, respectively), characterized by the presence of traces from different biofluids, suggesting a possible contamination through the patient right hand. Conversely, sample YH1_12 displayed microbiota traces mainly restricted to nasopharyngeal secretions and with a different biodiversity pattern (Shannon index 2.161), suggesting a possible contact of the stethoscope on the patient chest (probably contaminated after a sneeze without keeping the mask). Eventually, NGS provided a more comprehensive overview of all the environmental bacteria present on the sampled surfaces, confirming also those coming from the microbiota of human biological fluids.

**Figure 2.**
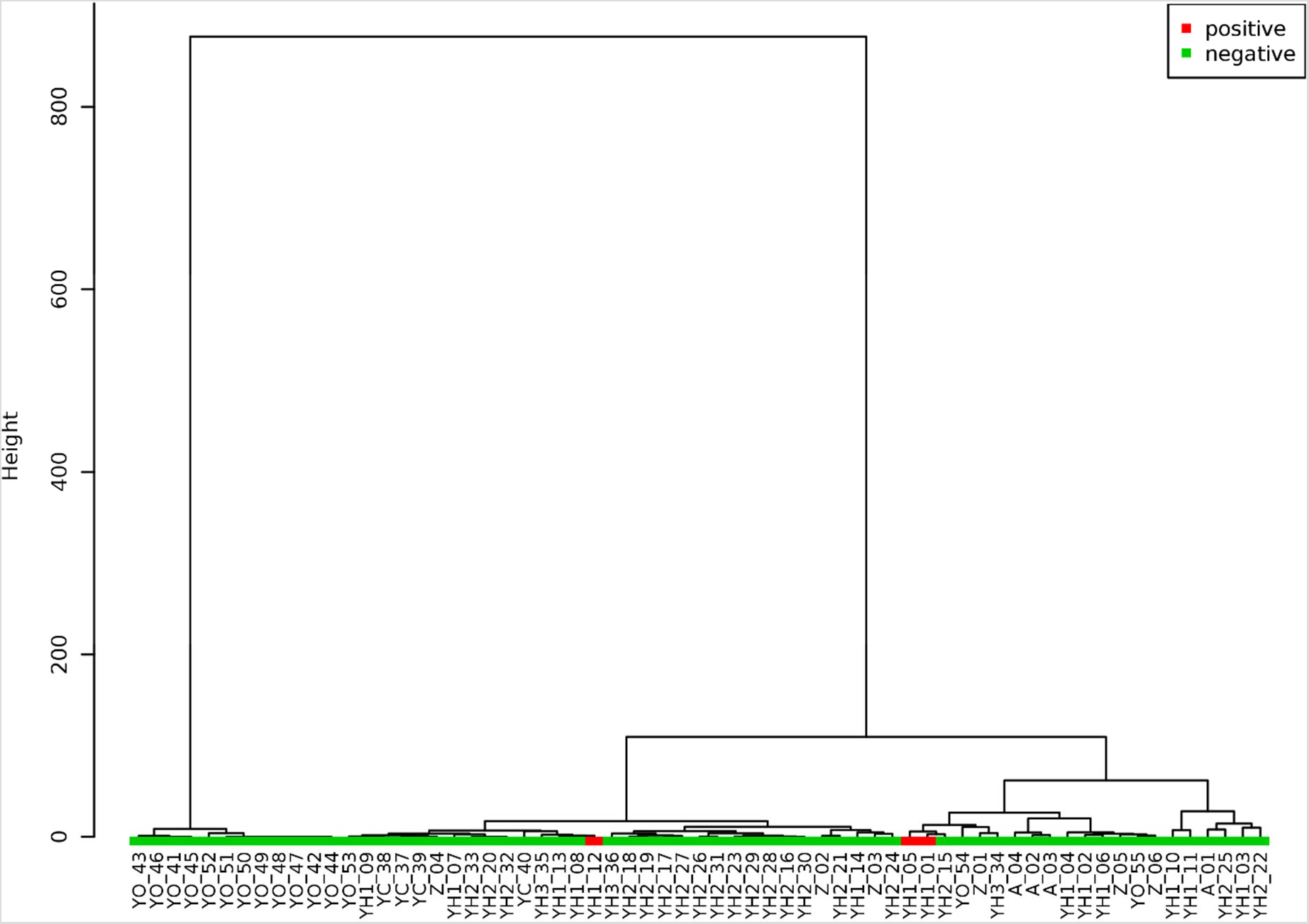
Hierarchical cluster analysis on Real Time PCR results. SARS-CoV-2 positive and negative samples are indicated in green and red, respectively. The hierarchical cluster was performed on raw CT data by Euclidean distance and Ward’s linkage (clustering to minimize the sum of squares of any two clusters).

**Figure 3.**
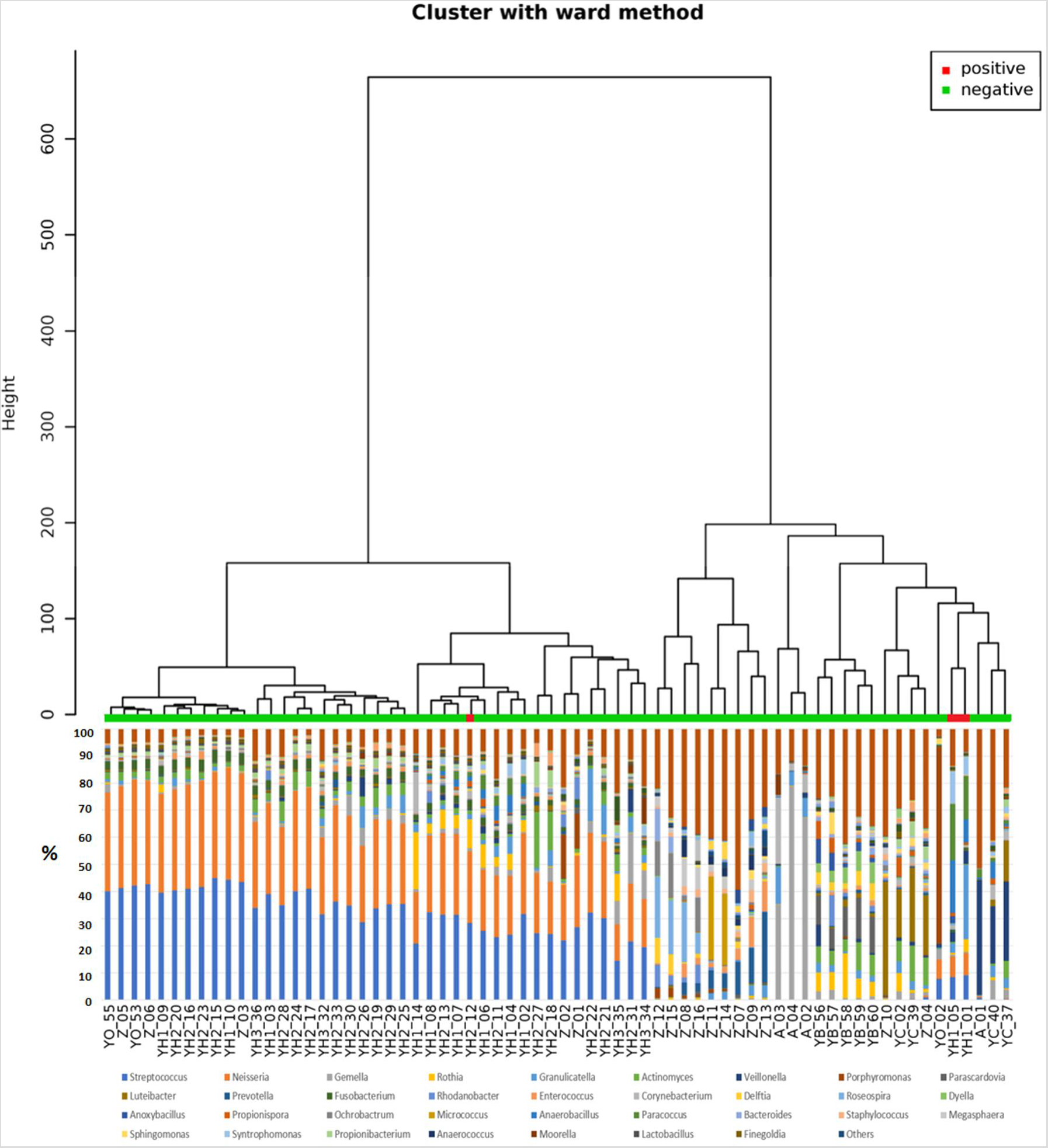
Hierarchical Clustering Dendrogram on 16S amplicon sequencing data. Dendrogram shows a hierarchical clustering of samples based on genus-level classifications. The bar chart under each sample summarizes the relative abundance of its genus-level classifications. In this analysis were included also environmental samples from playgrounds (Z_07-16) and indoor air (YB_56–60), without major anthropic contaminations. SARS-CoV-2 negative and positive samples are indicated in green and red, respectively.

**Figure 4.**
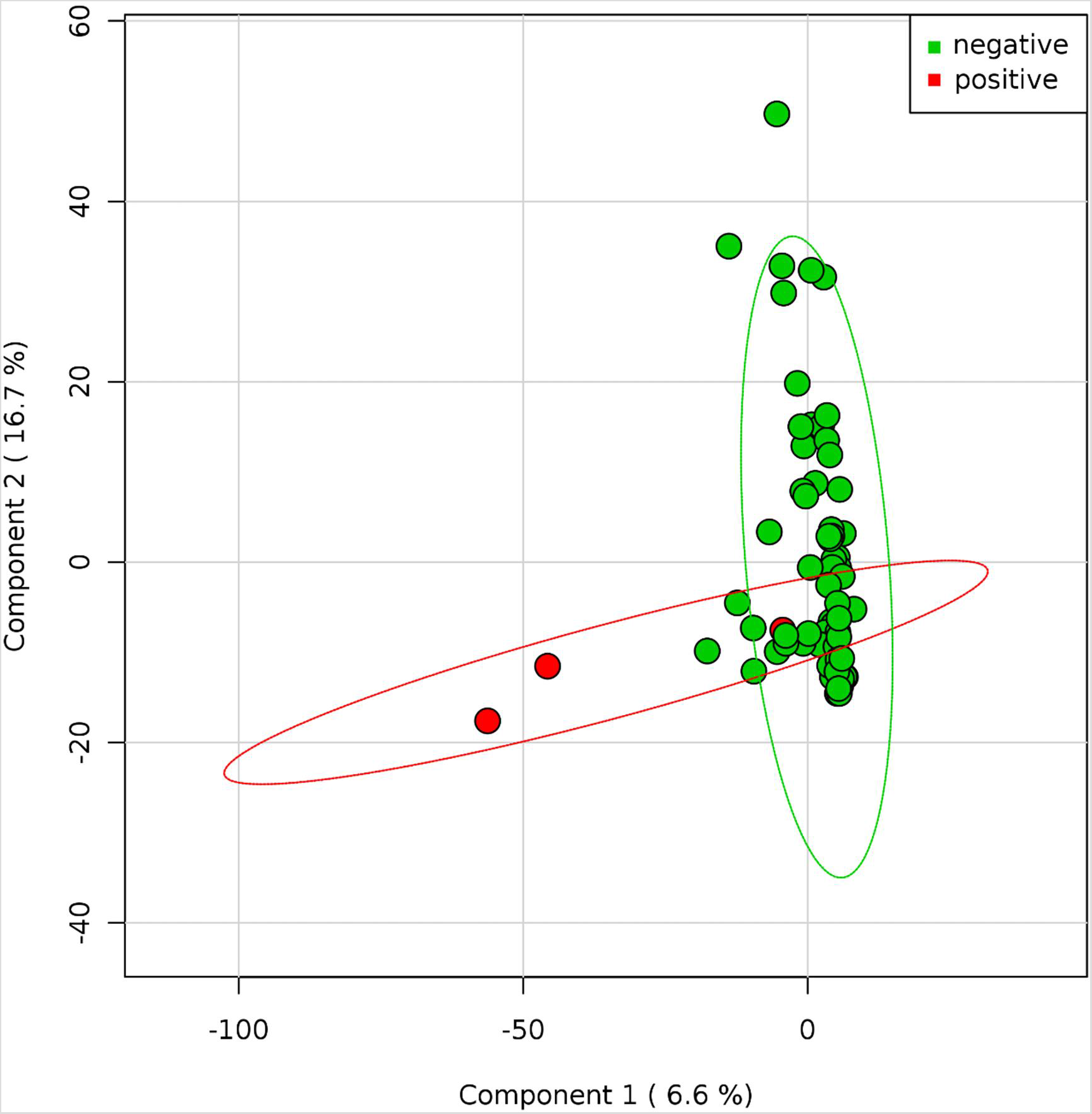
SARS-CoV-2 positive and negative samples. Principal Coordinate Analysis of the normalized relative abundance of all samples divided by negative and positive results of SARSCOV2. Data are plotted at the genus-level classification. The variance is explained for 6.6% and 16.7%, respectively for Component 1 and 2. SARS-CoV-2 positive (red) and negative (green) samples.

### 3.4 Fomites and environmental microflora

Environmental contamination through droplets or biofluids that can convey SARS-CoV-2 represents an additional component of the complex microflora detectable on a surface. Indeed, the identification of fomites by RT-PCR analysis of selected indicators emphasizes a very specific - and often minority-component of the resident or ectopic microflora. The analysis of the dataset by NGS showed not only the wide presence of fomites on several surfaces exposed to anthropic contamination, but also the inhabiting microorganisms or those from other environmental sources.

Samples collected from indoor and outdoor surfaces gathered independently from human biofluids (Figure 5). Within the outdoor group, those with a higher anthropic contamination overlapped with indoor samples, far from those where the environmental component was overwhelming. Interestingly, only an outdoor sample segregated outside –and in between- of both groups: the external handle of the entrance of a public building (Z_04), characterized by multiple contaminations of anthropic origin overlaying the outside microflora. Other indoor samples grouped together. These findings support the utility of microbiota data for tracing fomites in environmental samples and can sustain risk assessment for an indirect transmission of COVID-19. Nevertheless, the detection of traces of biological fluids in several environmental surfaces did not predict the presence of SARS-CoV-2, unless in virus positive points, even if sampling occurred during the pandemic period and in hospitals where COVID-19 patients were treated. Therefore, if fomites represent a risk themselves, the possibility for a contagion relays on the presence of the specific pathogen and its viability, satisfying the principles from the ancient Koch’s postulates (Segre, 2013). Moreover, even if detectable through its RNA, the environmental survival of SARS-CoV-2 depends on different indoor and outdoor factors, including sanitation, time from the release of the biological fluid, exposure to agents such as humidity, temperature, air circulation and sunlight (Aboubakr, 2020; Morawska, 2020; Yolitz, 2020; Ratnesar-Shumate, 2020; Biryukov, 2020). Therefore, viral infectivity can vary significantly between different environments and detection of fomites should be considered more as a putative indicator of transmission risk levels, than a danger itself. Monitoring droplets and biofluids by RT-PCR can help to prevent SARS-CoV-2 transmission by improving environmental surveillance and enforcing hygiene and sanitization procedures.

**Figure 5.**
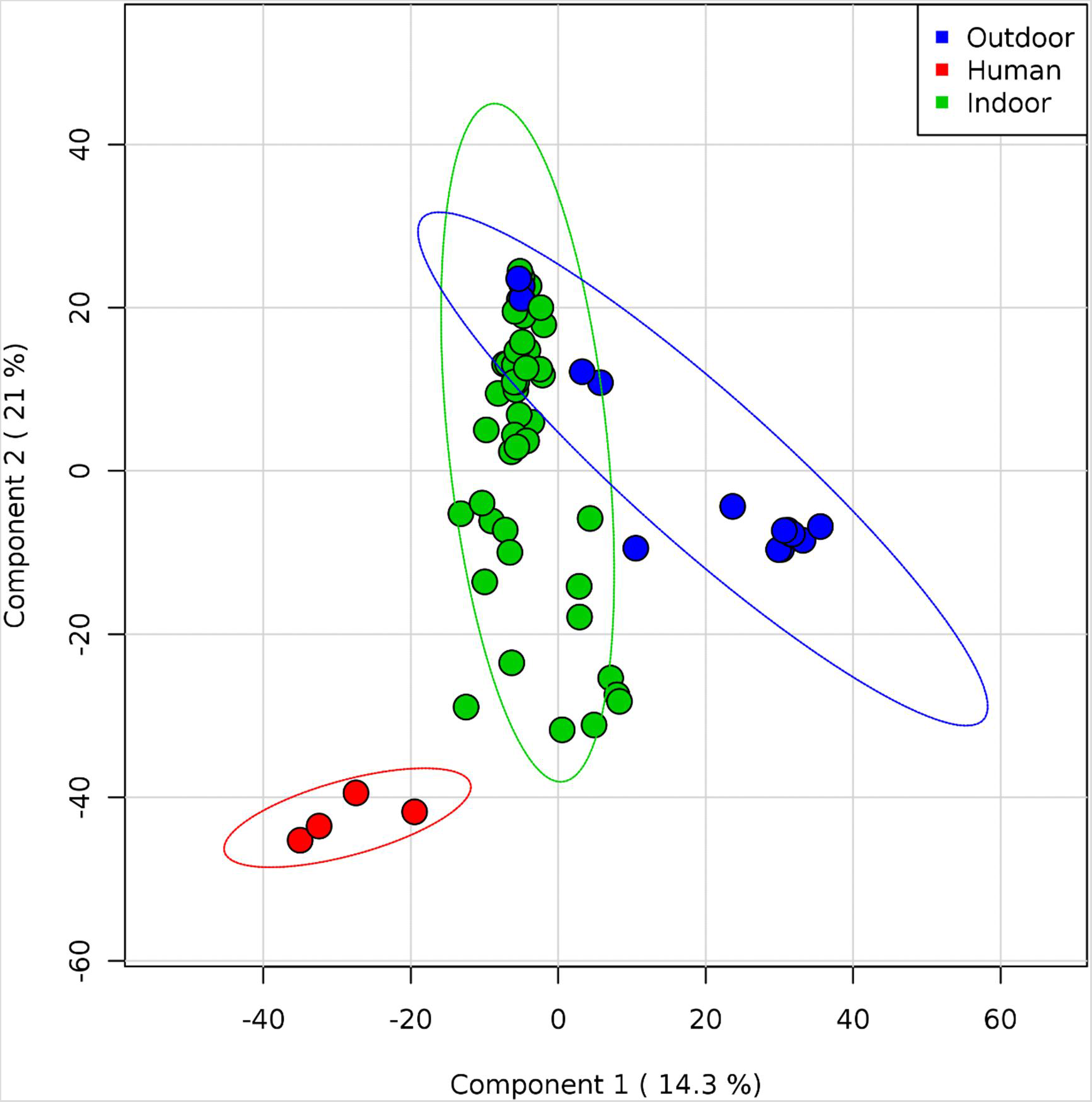
Whole microflora analysis of indoor and outdoor samples. Partial least square-discriminant analysis (PLS-DA) shows Pearson distance between different samples using phylogeny distribution based on 16S rRNA genes. Samples are colored according to the sampling point (red: human droplets; green: indoor; blue: outdoor). The variance is explained for 14.3 % and 21%, respectively for Component 1 and 2. Outdoor samples overlapping indoor samples are characterized by fomites, whereas the blue sample between groups is the outdoor handle of a building main entrance. All samples without a major presence of environmental microflora, but characterized by a prevalence of human microbiota from droplets biofluids, tend to segregate independently.

### 3.5 Critical issues and limits of the study

Even if several surfaces at risk of indirect transmission were evaluated, the study aim was not the quantification of the infectious risk. Comparisons between sampling points or buildings cannot be performed, not only for the sample size but also for the random collection from areas with a different incidence of disease. Indeed, different Italian regions were selected to avoid geographical bias, but different epidemiological burdens can affect the generalizability of the results. Therefore, we have simply reported an environmental scenario, suggesting a possible strategy to assess contaminations at risk for indirect transmission of COVID-19. Interestingly, SARS-CoV-2 positive samples were collected only in Emilia-Romagna, a region with the highest number of cases in Italy (Veronesi et al., 2020). Thus, this is not an epidemiological study aimed at comparing geographical areas or quantify a risk for a specific environment or sampled surface. Nevertheless, observed data clearly identify surfaces at risk and confirm the fundamental role played by hands. Regarding the chosen approach, RT-PCR is faster than NGS (3-5 hours vs 5-10 days), but still is not instantaneous, as other strategies aimed to detect contaminations on surfaces or medical devices (Valeriani, 2018 a; Lee, 2020).Tracing droplets and identifying biological fluids by RT-PCR or NGS is specific respect to other methods based on finding organic matter without a recognition of their origin, e.g. from human secretions rather than human cells, plant, animal or bacterial debris. Therefore, detection of droplets by RT-PCR is not a generic marker of hygiene but can find specific application also in assessing environmental risks for other communicable diseases following an indirect rout of transmission, such as flu-like infections (Otter et al., 2016; Petersen, 2020; Wei et al., 2020).

Another limitation concerns the set of experimental conditions for RT-PCR and NGS. Primers and probes for selected bacterial genes from selected bacterial markers where chosen because of their feasibility and effectiveness, but we did not made comparisons with different sequences, indicators or reaction conditions for RT-PCR. The same concern can be raised for the NGS approach, which was adopted for the analysis of 16S rDNA amplicon sequencing, following standard protocols. A whole genome analysis would have been more informative. However, it would have also been more expensive for materials and bioinformatic analysis, being less appropriate for public health surveys on a larger scale. Finally, we used arbitrary thresholds to quantify droplets contamination based on CT values, proposing the highest sensibility for droplets detection. A lower threshold would have provided more specific data, or it could have been adapted to the different kind of transmission risks or expected hygiene levels in the different environments.

## Conclusions

Environmental monitoring of fomites with a potential role in COVID-19 transmission, can be performed by RT-PCR. The general principle is based on the identification of anthropic contaminations by detecting their microbiota component.

Droplets and biological fluids were observed in most of the indoor places exposed to human presence, and on those surfaces frequently touched by hands. SARS-CoV-2 was not detectable diffusely in the environment, except in the proximity of an infected patient and in consistent association with fomites. The whole of the results supports the key role of handwashing and environmental sanitation in reducing risks related to indirect transmission of COVID-19 in hospitals or public areas, both indoor and outdoor. It also highlights the role of education and awareness in protecting the health of all.

In addition to searching for SARS-CoV-2 in the environment, the possibility to detect fomites by RT-PCR may provide a novel indicator for monitoring indirect transmission risks of COVID-19 as well as other communicable diseases transmitted through droplets, such as flu or flu-like infections.

## Data Availability

All data are reported in the manuscript. additional data can be required to the corresponding.

## Acknowledgments

The authors would like to thank dr. Elena Scaramucci, Tiziana Zilli and Pietro Robert for supporting bibliography search and management; Alessandro Stabile for assuring laboratory safety and acceptance of reagents during summertime; the Copan Italia S.p.A. Brescia Italy for providing eNATand UTM collection kits. This study was partially supported by University of Foro Italico Project (CDR2.TER012013) assigned to V.R.S.

